# Assessment of Accuracy and Safety of LabTest Checker (LTC-AI)

**DOI:** 10.1101/2023.09.26.23296157

**Authors:** Dawid Szumilas, Anna Ochmann, Katarzyna A. Zięba, Bartłomiej Bartoszewicz, Anna Kubrak, Sebastian Makuch, Siddarth Agrawal, Grzegorz Mazur, Jerzy Chudek

## Abstract

**Background:** In recent years, the implementation of artificial intelligence (AI) in healthcare is progressively transforming medical fields, with Clinical Decision Support Systems (CDSS) as a notable application. Laboratory tests are vital for accurate diagnoses, but their increasing reliance presents challenges. The need for effective strategies for managing laboratory test interpretation is evident from the millions of monthly searches on test results’ significance. The potential role of CDSS in laboratory diagnostics gains significance, however, more research needs to explore this area.

**Objective:** The primary objective of our study was to assess the accuracy and safety of LabTest Checker (LTC), a CDSS designed to support medical diagnoses by analyzing both laboratory test results and patients’ medical histories

**Methods:** This cohort study embraced a prospective data collection approach. A total of 101 patients were enrolled, aged 18 and above, in stable condition, requiring comprehensive diagnosis. A panel of blood laboratory tests was conducted for each participant. Participants utilized LabTest Checker for test result interpretation. Accuracy and safety of the tool were assessed by comparing AI-generated suggestions to experienced doctor (consultant) recommendations, considered the gold standard.

**Results:** The system achieved a 74.3% accuracy and 100% sensitivity for emergency safety and 92.3% sensitivity for urgent cases. It potentially reduced unnecessary medical visits by 41.6% and achieved an 82.9% accuracy in identifying underlying pathologies.

**Conclusion:** This study underscores the transformative potential of AI-based CDSS in laboratory diagnostics, contributing to enhanced patient care, efficient healthcare systems, and improved medical outcomes. LabTest Checker’s performance evaluation highlights the advancements in AI’s role in laboratory medicine.

## 1. Introduction

In recent times, the implementation of artificial intelligence (AI) within diverse medical domains has garnered significant attention and practical application [1]. AI-driven technology has sparked a transformative wave in healthcare, introducing inventive solutions to enhance patient care, diagnosis, and decision-making processes [2]. A notable instance of AI’s application is evident in the emergence of Clinical Decision Support Systems (CDSS), direct tools designed to streamline healthcare decision-making [3].

Laboratory tests are essential in modern healthcare, providing valuable insight into a patient’s health status and improving the accuracy of diagnosing medical conditions. Interpretation of laboratory test results is a complex process requiring medical expertise and knowledge. However, the mounting reliance on laboratory testing poses a formidable challenge for healthcare systems, particularly in regions where tests are often administered without direct medical oversight, as seen in Poland [4].

The significance of this challenge is highlighted by the substantial volume of inquiries related to laboratory test result interpretation. Data indicates that, in Poland alone, there are approximately 7 million monthly searches concerning the significance of laboratory test results. On a larger scale, within the European Union (EU), this number escalates to around 82 million monthly searches based on data from SENUTO and Google AdWords [5]. These cases emphasize the need for effective strategies to manage laboratory test interpretation in modern healthcare settings.

Given the widespread use of laboratory diagnostics and the inherent complexities tied to test result interpretation, there is a growing interest in exploring the potential of CDSS within this realm. The efficacy and safety of CDSS have been demonstrated in various medical contexts, such as symptom assessment tools [6,7]. Even so, more research has delved into integrating CDSS into laboratory diagnostics.

This introduction aims to shed light on the underexplored area of AI-based CDSS in laboratory diagnostics. By exploring the potential benefits, challenges, and implications of implementing CDSS in this domain, we aim to bridge the gap between AI advancements and laboratory medicine. A comprehensive grasp of CDSS’s role in laboratory diagnostics stands poised to reshape healthcare delivery and improve patient outcomes and healthcare system efficiency—particularly in regions where tests are frequently conducted without direct medical supervision.

Several studies have assessed the effectiveness and safety of AI-driven symptom checkers, tools designed to aid patients in self-diagnosing symptoms and making informed healthcare choices [8–10]. These tools employ algorithms and databases to generate potential diagnoses based on user inputs.

A notable study conducted by Semigran et al. scrutinized the diagnostic precision of 23 distinct symptom checkers, comparing their outcomes against physician diagnoses [11]. The investigation disclosed that symptom checkers achieved accurate diagnoses in 34% of instances, while physicians achieved 58% accuracy. Despite relatively lower accuracy, the study underscored the potential of symptom checkers in offering reasonable differential diagnoses and supporting patient decision-making.

A more recent study by Hennemann al. (2022) evaluated the performance of an app-based symptom checker within the realm of mental disorders [12]. Results revealed that the studied symptom checker demonstrated moderate-to-good accuracy in suggesting conditions for mental disorders concerning formal diagnosis, albeit with variations across disorder categories and interrater reliability. The symptom checker’s primary condition suggestion corresponded with interview-based diagnoses in 51% (25/49) of cases, with at least one of the initial five condition suggestions aligning in 69% (34/49) of cases across the patient cohort. Accuracy fluctuated across disorder categories, ranging from 82% precision for somatoform and related disorders, 65% for affective disorders, to 53% for anxiety disorders. The study concluded that symptom checkers hold promise as supplementary screening tools in the diagnostic process. Still, their diagnostic efficacy requires assessment in more extensive samples and comparison with alternative diagnostic methods.

This paper addresses the current status of AI-based technologies in healthcare, specifically focusing on implementing CDSS in direct-to-patient tools. After emphasizing the importance of laboratory diagnostics in contemporary healthcare and the challenges tied to test result interpretation, we examine the existing but limited literature concerning CDSS’s role in laboratory diagnostics, underscoring the need for further research and advancement in this domain. The objective of this study is to evaluate the performance of a novel clinical decision support system named ‘LabTest Checker’ in a cohort of adult patients requiring laboratory testing. The main question it aims to answer pertains to the accuracy and safety of LabTest Checker.

## 2. Materials and Methods

### 2.1. Description of LTC technology

LabTest Checker (LTC) is an intricate medical software designed to provide assistance in the preliminary medical diagnosis process through the analysis of laboratory test results and comprehensive medical history. By leveraging advanced algorithms and data analytics, LTC empowers patients and healthcare practitioners to derive insightful conclusions and make informed decisions. Beyond its analysis of lab results, LTC augments its diagnostic prowess by engaging patients in a series of meticulously crafted inquiries concerning medical history, symptoms, and pertinent risk factors. This integration equips LTC to pinpoint potential ailments and offer tailored guidance in further diagnostic and therapeutic processes. This innovative tool effectively evaluates an individual’s health status and detects potential medical issues by merging lab test findings with the patient’s medical history. Through this methodical scrutiny and correlation of pivotal data, LTC empowers patients and healthcare providers to establish more accurate diagnoses, thus elevating patient care and outcomes.

### 2.2. Patient recruitment

This cohort study embraced a prospective data collection approach. A total of 101 patients aged 18 and above, in stable condition but requiring comprehensive diagnosis, were enrolled. Comprehensive diagnosis refers to cases where diagnosis based solely on subjective evaluation and physical examination is unattainable, necessitating in-depth assessment through laboratory tests. Inclusion criteria encompassed: a) age above 18 years, b) requirement of in-depth laboratory test investigation. The only exclusion criterion was pregnancy.

### 2.3. Study design

*A panel of blood laboratory tests, including a lipid profile, ESR (erythrocyte sedimentation rate), hs-CRP (high sensitive C-reactive protein), creatinine, urea, iron, liver enzymes (ALT, AST, GGT), sodium, potassium, glucose, uric acid, thyroid-stimulating hormone (TSH), and complete blood count, was conducted for each participant. Participants utilized LabTest Checker for test result interpretation, which was then compared to an internal medicine specialist’s interpretation to evaluate tool accuracy and safety*.

Patients presenting at the Emergency Department underwent laboratory tests and provided health-related information under a doctor’s supervision. This encompassed biometric details, medical history, medications, substances used, family history, symptoms, and prior test results. Based on this data and test outcomes, AI algorithms suggested underlying pathology and diagnostic-therapeutic guidance.

Accuracy and safety were assessed by comparing AI-generated suggestions to experienced doctor (consultant) recommendations, considered the gold standard. The consultant, blinded to LTC results, categorized the urgency of physician interaction for each test (emergency, urgency, routine, self-care) (Table 1). Following assessment, LTC results were disclosed to the consultant to evaluate if adhering to LTC recommendations could avoid needless medical visits and whether LTC accurately identified potential result deviations’ causes.

**Table 1.** Diagnostic and therapeutic recommendations generated by LTC and specialist recommendations were categorized to assess LTC’s precision. Sensitivity for the emergency category was computed as the ratio of LTC’s correct emergency identifications to the physician’s emergency identifications = A / (A + B + C + D). Similarly, sensitivity for the urgency category was calculated as F / (E + F + G + H). Triage accuracy = (A + F + L + R) / total patients in the study. Triage safety = (A + E + F + J + K + L + N + O + P + R) / total patients in the study.

|  |  | Urgency category of contact with doctor, assigned by LTC |  |  |  |
| --- | --- | --- | --- | --- | --- |
|  |  | emergency | urgency | routine | self-care |
| Urgency category of contact with doctor, assigned by consultant | emergency | A | B | C | D |
|  | urgency | E | F | G | H |
|  | routine | J | K | L | M |
|  | self-care | N | O | P | R |

Owing to the technology’s design, certain variables were excluded from determining pathology identification accuracy:

a. Interpretations labeled as urgent or requiring immediate contact with a doctor were omitted. In emergency and urgent contexts, patient safety mandates minimal questioning by the technology. Subsequent actions (diagnostics, potential treatment) are to be conducted by a doctor, with the patient’s prompt contact being of utmost importance. During urgent situations, triage takes precedence over identifying pathology causes.
b. Interpretations categorized as “end of diagnostic - no need for doctor contact” were omitted. This pertains to valid results or insignificant deviations from the norm that do not signify pathology.

The study was approved by the Bioethics Committee of the Medical University of Silesia (Approval Code: PCN/CBN/0052/KB1/115/I/22, Approval Date: 08.11.2022).

### 2.4. Statistics

A power analysis was performed to determine the statistical power of this study, considering a total sample size of 101 participants in a single group of patients, which was predetermined in the study design. The power analysis was conducted using the G*Power software (version 3.1.9.7). The power analysis was based on a one-tailed test with an alpha level of 0.05. The effect size was calculated at 0.36. Using these parameters and the total sample size of 101, the power analysis indicated that the study would have moderate statistical power to detect a significant effect size within a single group of patients. The estimated power achieved with the given sample size was 0.82, indicating that the study had a reasonable likelihood of detecting meaningful differences within the group.

Outcomes measures were pre-specified and calculated with 95% confidence intervals. Wilson score method was used to produce confidence intervals for sensitivity to emergency, sensitivity to urgency, accuracy of triage, safety of triage, and reduction of unnecessary visits. Calculations were performed using the statistical software package *Statistica* 13.0 PL *(TIBCO Software Inc*., *Palo Alto, CA, U*.*S*.). Analytic data is presented as point estimates and 95% confidence intervals (±95% CI). A p-value of < 0.05 was considered to be significant.

## 3. Results

In the context of this study, the triage accuracy in 101 patients cohort attained 74.3%, with a safety sensitivity of 100% for identifying emergency cases and a sensitivity of 92.3% for detecting urgent cases. Implementation of the system led to a noteworthy 41.6% reduction in unnecessary medical visits, and its accuracy in identifying the underlying pathology achieved 82.9%.

The system classified patients based on urgency: 9 patients required immediate contact, 41 needed urgent contact, 50 warranted routine contact, and 1 patient did not necessitate doctor contact, falling into the self-care category. Analysis by the consultant revealed disparities in urgency category assignments for 26 patients. Notably, the technology overestimated urgency for 25 patients, including cases where the consultant recommended urgent contact, but the technology indicated immediate or scheduled contact. However, the technology inaccurately assessed the urgency for one patient, failing to align with the specialist’s urgent contact suggestion, instead proposing scheduled contact. These findings collectively underscore the triage system’s effective urgency categorization while also pinpointing areas for enhancement to improve precision, diminish disparities, and prevent false negatives. These findings are detailed in Table 2.

**Table 2.** Classification outcomes of diagnostic-therapeutic recommendations proposed by LTC and those provided by the specialist.

|  |  | Urgency category of contact with doctor, assigned by LTC |  |  |  |
| --- | --- | --- | --- | --- | --- |
|  |  | Emergency | Urgency | Routine | Self-care |
| Urgency category of contact with doctor, assigned by consultant | Emergency | 7 |  |  |  |
|  | Urgency | 1 | 24 | 1 |  |
|  | Routine | 1 | 15 | 43 |  |
|  | Self-care |  | 2 | 6 | 1 |

## 4. Discussion

The promising results obtained from the evaluation of LabTest Checker (LTC) showcase the potential of AI-driven tools in assisting patients and medical professionals in navigating the complexities of laboratory test result interpretation. An accuracy rate of 74.3% showcases LTC’s capability to furnish dependable medical recommendations grounded in blood test results, a development that holds promise for enhancing operational efficiency in the medical domain. Particularly noteworthy is LTC’s impressive safety sensitivity of 100% for identifying emergency cases and a high sensitivity of 92.3% for detecting urgent cases. These results imply the system’s adeptness in identifying critical scenarios, aligning with its intended role of providing secure and precise medical counsel.

However, it’s important to note that due to the recent emergence of AI-powered CDSS technologies, there is a dearth of extensive research in this realm. One such study by Gräf et al. compared physician and AI-based symptom checker diagnostic accuracy, where the AI achieved a diagnostic accuracy of 70% [13]. Furthermore, a systematic review of 10 studies revealed consistently low diagnostic accuracy (range: 19–37.9%), while triage accuracy (range: 48.8–90.1%) was relatively higher but displayed variability among different symptom checkers [9]. This underscores the necessity for more robust research efforts and improved regulatory measures in this evolving field.

While the study yielded promising results, several inherent limitations should be acknowledged when assessing the accuracy and safety of LTC. Firstly, the sample size was relatively small, comprising only 101 participants. Although efforts were taken to ensure analytical strength, a larger and more diverse sample would enhance the generalizability of findings to the broader population. Another constraint lies in the difference between the participant sample and LTC’s intended target population. Study participants were drawn from an emergency room environment, whereas the technology is meant to be used independently by individuals at home without immediate medical guidance. Such variations might influence the technology’s performance and should be considered while interpreting results. Furthermore, the study allowed participants to seek guidance from medical professionals when faced with uncertainties while filling out the questionnaire, which might not mirror real-world usage where such guidance might not be readily accessible. While this provision was aimed at optimizing data quality, it could have potentially introduced an artificial element, warranting caution when considering the practical implications of the technology’s recommendations. These limitations underscore the necessity for future research involving more representative samples and real-world usage scenarios to validate the robustness and effectiveness of emerging CDSS technologies. By exploring the intersection of AI and laboratory diagnostics, we aim to lay the groundwork for future progress and foster a deeper comprehension of AI-based CDSS potential in reshaping laboratory medicine.

## 5. Conclusions

Our study has shown the potential impact of integrating AI into laboratory diagnostics *via* the LabTest Checker software. The findings underscore the promise of AI-driven Clinical Decision Support Systems in enhancing healthcare decision-making. The attainment of a high accuracy rate alongside notable safety sensitivity underscores the system’s ability to precisely identify medical conditions, facilitating optimal advice grounded in various factors encompassing blood testing and patient medical history.

As the landscape of AI-assisted healthcare evolves, this study contributes to the ongoing discourse regarding AI’s role in medical diagnostics. It emphasizes the necessity for comprehensive research bridging the gap between AI technologies and laboratory medicine. While the results exhibit encouraging potential, they also pinpoint areas requiring further exploration.

## Data Availability

All data produced in the present study are available upon reasonable request to the authors

## Informed Consent Statement

All patients provided written informed consent before undergoing screening for study eligibility.

## Data Availability Statement

All data produced in the present study are available upon reasonable request to the authors

## Acknowledgments

We would like to acknowledge Karol Dobrzyński for his valuable assistance in preparing this manuscript.

## References

1. Davenport, T.; Kalakota, R. The potential for artificial intelligence in healthcare. Futur. Healthc. J. 2019, 6, 94–98, doi:10.7861/futurehosp.6-2-94.

2. Johnson, K.B.; Wei, W.Q.; Weeraratne, D.; Frisse, M.E.; Misulis, K.; Rhee, K.; Zhao, J.; Snowdon, J.L. Precision Medicine, AI, and the Future of Personalized Health Care. Clin. Transl. Sci. 2021, 14, 86–93, doi:10.1111/cts.12884.

3. Castaneda, C.; Nalley, K.; Mannion, C.; Bhattacharyya, P.; Blake, P.; Pecora, A.; Goy, A.; Suh, K.S. Clinical decision support systems for improving diagnostic accuracy and achieving precision medicine. J. Clin. Bioinforma. 2015, 5, 1–16, doi:10.1186/s13336-015-0019-3.

4. Polacy leczą się sami. 90 proc. zażywa leki bez recepty | WP abcZdrowie Available online: https://portal.abczdrowie.pl/polacy-lecza-sie-sami-90-proc-zazywa-leki-bez-recepty (accessed on Aug 18, 2023).

5. Polskie AI ma w kilka sekund zweryfikować wyniki badań. Wszystko online i bez wychodzenia z domu Available online: https://bizblog.spidersweb.pl/analiza-wynikow-badan-online (accessed on Aug 18, 2023).

6. Fraser, H.; Coiera, E.; Wong, D. Safety of patient-facing digital symptom checkers. Lancet 2018, 392, 2263–2264, doi:10.1016/S0140-6736(18)32819-8.

7. Sutton, R.T.; Pincock, D.; Baumgart, D.C.; Sadowski, D.C.; Fedorak, R.N.; Kroeker, K.I. An overview of clinical decision support systems: benefits, risks, and strategies for success. npj Digit. Med. 2020, 3, 1–10, doi:10.1038/s41746-020-0221-y.

8. Chambers, D.; Cantrell, A.J.; Johnson, M.; Preston, L.; Baxter, S.K.; Booth, A.; Turner, J. Digital and online symptom checkers and health assessment/triage services for urgent health problems: Systematic review. BMJ Open 2019, 9, e027743, doi:10.1136/bmjopen-2018-027743.

9. Wallace, W.; Chan, C.; Chidambaram, S.; Hanna, L.; Iqbal, F.M.; Acharya, A.; Normahani, P.; Ashrafian, H.; Markar, S.R.; Sounderajah, V.; et al. The diagnostic and triage accuracy of digital and online symptom checker tools: a systematic review. npj Digit. Med. 2022, 5, doi:10.1038/s41746-022-00667-w.

10. Nateqi, J.; Lin, S.; Krobath, H.; Gruarin, S.; Lutz, T.; Dvorak, T.; Gruschina, A.; Ortner, R. From symptom to diagnosis—symptom checkers re-evaluated: Are symptom checkers finally sufficient and accurate to use? An update from the ENT perspective. HNO 2019, 67, 334–342, doi:10.1007/s00106-019-0666-y.

11. Semigran, H.L.; Linder, J.A.; Gidengil, C.; Mehrotra, A. Evaluation of symptom checkers for self diagnosis and triage: Audit study. BMJ 2015, 351, doi:10.1136/bmj.h3480.

12. Hennemann, S.; Kuhn, S.; Witthöft, M.; Jungmann, S.M. Diagnostic Performance of an App-Based Symptom Checker in Mental Disorders: Comparative Study in Psychotherapy Outpatients. JMIR Ment. Heal. 2022, 9, e32832, doi:10.2196/32832.

13. Gräf, M.; Knitza, J.; Leipe, J.; Krusche, M.; Welcker, M.; Kuhn, S.; Mucke, J.; Hueber, A.J.; Hornig, J.; Klemm, P.; et al. Comparison of physician and artificial intelligence-based symptom checker diagnostic accuracy. Rheumatol. Int. 2022, 42, 2167–2176, doi:10.1007/s00296-022-05202-4.

